# Internal and External Protective Factors Associated with the Secondary Traumatic Stress Component of Compassion Fatigue, in Feral Cat Caregivers

**DOI:** 10.64898/2026.03.05.26347725

**Authors:** Cristina Costa-Santos, Rui Vidal, Sara Lisboa, Paulo Vieira-de-Castro, Ana Monteiro, Ivone Duarte

## Abstract

Compassion fatigue is a well-documented hazard among healthcare and veterinary professionals, yet the psychological toll on informal caregivers of feral cat colonies, likely numbering several tens of thousands in Portugal, remains largely unexplored. This cross-sectional study examines internal and external factors associated with the secondary traumatic stress component of compassion fatigue among 172 informal caregivers in Portugal. Secondary traumatic stress refers to work-related secondary exposure to individuals who have experienced extremely stressful or traumatic events. Structured telephone interviews assessed sociodemographics, colony management, compassion satisfaction, resilience, spiritual well-being, and perceived social support. Univariate and multivariable linear regression identified predictors of secondary traumatic stress. Results indicate that 47% of participants experienced moderate secondary traumatic stress, and 10% reported high levels. Multivariable analysis revealed that caring for large colonies (more than 25 cats) and being unemployed were significantly associated with higher fatigue. Conversely, older age, higher perceived family support, and the resilience dimension of serenity served as protective factors. Interestingly, finding meaning in life was positively correlated with fatigue, suggesting that caregivers who perceive their role as central to their life purpose may become more emotionally invested, increasing vulnerability to distress when unable to help animals. Official colony registration and formal institutional support did not significantly alleviate fatigue. These findings highlight that institutional support alone is insufficient to mitigate fatigue among informal caregivers, who experience significant distress driven by both practical burdens and profound emotional involvement. The most frequently reported concern among caregivers was the inability to cover the costs of feeding and veterinary care for the cats. Interventions must address both external needs (e.g., support to cover veterinary and feeding expenses for the cats) and internal coping mechanisms. Implementing psychosocial support alongside trap-neuter-return programs may also improve caregiver well-being and foster sustainable urban feral cat management. This underscores a One Health perspective, demonstrating that animal health is closely interconnected with human well-being and environmental health.

## Introduction

Compassion satisfaction is a degree of fulfillment, sense of purpose and satisfaction derived from working as a care provider [1]. Compassion stress, on the other hand, is the inevitable psychological distress that arises from assisting others in distress or facing danger. Compassion fatigue is a condition of exhaustion accompanied by biological, physiological, and emotional dysfunction caused by extended exposure to compassion stress [2]. In recent years, the term compassion fatigue has emerged in the literature to describe a condition specific to professionals in helping contexts (healthcare professionals, teachers, police officers), who are in direct contact with the suffering of others [3]. The term compassion fatigue is widely used in the scientific literature and has been recognized since 2016 as a MeSH term in MEDLINE, defined as “emotional distress caused by repeated or prolonged expression of compassion or empathy. It may occur in individuals working in caregiving professions.” In MEDLINE, the term is treated as synonymous with secondary traumatic stress. However, this terminology is not universally accepted. Some scholars argue that compassion entails a prosocial motivation to alleviate suffering, whereas what many caregivers experience may be more accurately described as empathic distress or empathic strain, a state arising from emotional over-identification with others’ pain rather than from compassion itself [4]. Active empathy engages neural circuits associated with pain and suffering, whereas compassion activates regions related to positive affect and caregiving [5]. Moreover, according to the Professional Quality of Life (ProQoL) scale, compassion fatigue consists of two components. The first component, Burnout, refers to feelings such as exhaustion, frustration, anger, and depression that are typical of burnout. The second component, Secondary Traumatic Stress, involves negative feelings driven by fear and work-related trauma. In other words, this element of compassion fatigue reflects work-related secondary exposure to individuals who have experienced extremely stressful or traumatic events. It may include symptoms such as sleep difficulties, intrusive images, or avoidance of reminders of another person’s traumatic experiences. This component of compassion fatigue is closely related to vicarious trauma, as the two constructs share many similar characteristics [1].

A systematic review and meta-analysis demonstrated that nurses report moderate levels of compassion fatigue and satisfaction [6]. Interventions aimed at addressing compassion fatigue in nursing professionals have been shown to be effective. These interventions include individual-level strategies, such as mindfulness and resilience training, as well as organizational approaches focused on improving the work environment and professional support [7].

The concept of compassion fatigue or Secondary Traumatic Stress has only recently been extended to include those working with animals [8]. Veterinary careers often draw individuals with a strong sense of compassion, reflected in the joy and fulfillment they experience from helping and caring for others [9, 10]. However individuals working in the veterinary field are known to face significant challenges in terms of mental health, encompassing symptoms such as depression, anxiety, and stress [11]. The veterinary professionals seems to be at higher risk for suicide [12] and life satisfaction appears to reduce psychological distress mainly through internal mechanisms like compassion satisfaction and resilience, rather than external factors such as perceived social support [13].

Historically, cats were domesticated in early agricultural societies because they controlled rodents attracted to stored grain [14]. While cats may still retain this function in rural environments, in urban and suburban settings feral cat colonies rarely provide a direct practical benefit to caregivers. Instead, caregiving is primarily motivated by ethical and emotional concerns. Consequently, caregivers invest time, financial resources, and emotional effort in animals without utilitarian return, while remaining continuously exposed to illness, injury, and death within the colony. This shift from a functional to an affective human–animal relationship may increase vulnerability to compassion-related psychological distress [15]. A recent study conducted by the University of Aveiro in collaboration with the Portuguese Institute for Nature Conservation and Forests estimated that there are approximately 830,541 feral cats in Portugal. The estimate was derived from extrapolations based on modelling of survey data gathered through the first national survey of feral animals [16]. The presence of numerous intact feral cats in various areas of cities not only harms their well-being but also causes issues for residents, including reproduction, noise, odors, and unsanitary conditions. In response, several cities around the world implemented Trap-Neuter-Return (TNR) programs, a program used to control feral cat colonies by reducing the number of kittens being born. Several studies, conducted in different countries, have evaluated TNR as a method for managing feral cat colonies and have observed benefits in reducing colony size [17, 18]. In TNR programs, animal welfare organizations coordinate informal caregivers to implement management plans duly authorized by the municipal veterinarian. This ensures that the public space of the colony is cleaned and sanitized, free from waste or food remnants, to prevent the proliferation of pests. In Portugal, animal welfare organizations and the municipality coordinate the TNR management plan for an identified cat colony, while a voluntary caregiver is then responsible for the cats’ daily care. Although the way of working and the level of support provided to voluntary caregivers vary from one municipality to another, this is generally how the system operates. Furthermore, there are several feral cat colonies not managed by the municipality or animal protection associations that are also cared for daily by informal caregivers, spontaneously. These informal caregivers, who care for both municipally identified and unidentified colonies, play an essential role in improving animal well-being, reducing public health risks, and promoting a compassionate and effective approach to managing feline populations. Despite the importance of their work, these caregivers are often underappreciated and sometimes mistreated by neighbors.

The present problem can be framed within the One Health paradigm, which recognizes the interdependence between human, animal, and environmental health [19; 20]. In the context of feral cat colonies, these three dimensions do not merely coexist but are dynamically interconnected. The psychological well-being of informal caregivers constitutes an essential human health component within this system, as their sustained engagement directly influences animal welfare and environmental conditions in shared urban spaces. Compassion fatigue may compromise caregivers’ ability to maintain consistent feeding routines, adequate sanitation practices, and monitoring of animal health, thereby affecting both feline welfare and the sanitary conditions of public spaces. Conversely, unmanaged feral cats colonies, community conflicts, residents’ complaints, and repeated exposure to illness and death within the colony may intensify caregivers’ psychological distress. This reciprocal dynamic represents a One Health feedback loop, in which human psychological burden, animal welfare, and environmental health continuously influence one another [21]. Although One Health frameworks have traditionally emphasized zoonotic disease transmission and environmental risk mitigation [19], less attention has been given to the psychological sustainability of informal actors who sustain community-based animal management systems. Recognizing compassion fatigue as a One Health concern expands the paradigm to include caregiver mental health as a structural determinant of sustainable human–animal coexistence.To the best of our knowledge, while factors and mechanisms that reduce psychological distress have been studied among veterinary staff [12, 13], they have not been examined in the context of informal caregivers of feral cat colonies. We believe that these informal caregivers may experience as much or even higher levels of psychological distress than veterinary staff. In fact, they face similar adversities but often lack the formal training of veterinary professionals. Furthermore, their work is not as widely recognized or valued by society. Moreover, as far as we know, the factors that protect feral cat caregivers from compassion fatigue or secondary traumatic stress have not been studied.

In psychology, resilience refers to the capacity to respond positively to adverse events, with a view to promoting health, well-being, and quality of life. In essence, it represents the individual’s capacity to achieve positive adaptation and social functioning following an experience that could have been traumatic. Resilience can be divided into four factors: Perseverance: refers to the enthusiastic persistence in finding solutions to problems and overcoming adversity; Sense of Meaning in Life: concerns the awareness of having something meaningful to live for and the notion that life has a purpose on which the individual focuses, avoiding becoming fixated on issues that cannot be resolved; Serenity: denotes a balanced, purpose-focused perspective on one’s life, with the capacity to accept a variety of experiences (including adverse ones) in a serene and enthusiastic manner, and the ability to exercise self-esteem; Self-Sufficiency and Self-Confidence: captures a sense of uniqueness and the awareness that each person’s life trajectory is unique, with certain stages faced not in a group but in solitude, enabling the person to be self-reliant and essentially depend on themselves [22].

Spiritual well-being can be defined as a dynamic way of being that is reflected in the quality of the relationships an individual establishes across four dimensions: with oneself, with others, with the environment, and with someone who transcends the human domain. The personal dimension concerns how a person relates to themselves in terms of meaning, purpose, and life values. It presupposes self-knowledge and self-awareness. The community domain refers to the quality and depth of interpersonal relationships and includes feelings such as love, justice, and faith in humanity. The environmental domain encompasses relationships with the physical and biological world (caring for and protecting it), expressed through feelings of connection with nature. Finally, the transcendental domain refers to one’s relationship with someone beyond the human sphere and is expressed through worship or reverence toward the source of the universe’s mystery [23].

In this context of informal animal caregivers, the Secondary Traumatic Stress component of Compassion Fatigue can be more relevant rather than the Burnout component. In fact, Secondary Traumatic Stress captures the emotional distress associated with indirect exposure to others’ suffering [1], which is central to the experience of individuals caring for vulnerable or injured animals. In line with this perspective, previous findings indicate that individuals reporting higher levels of compassion fatigue are typically those who perceive themselves as being more frequently exposed to the suffering of those they care for, supporting Figley’s theory [3] that compassion fatigue is strongly related to prolonged exposure to Secondary Traumatic Stress [24].

This study aims to examine the influence of external factors, such as perceived social support of informal caregivers and feral cat colony characteristics, and internal mechanisms, including compassion satisfaction, resilience, and spiritual well-being, on the Secondary Traumatic Stress component of Compassion Fatigue of caregivers of feral cat colonies.

## Material and Methods

This is a cross-sectional study. The target population consists of informal caregivers of feral cat colonies, regardless of whether the colonies are formally recognized by local municipalities.Inclusion criteria included caring for stray cats, living in Portugal, and being a Portuguese speaker.

Data will be collected through structured telephone interviews conducted after obtaining informed consent. Recruitment will rely on word-of-mouth dissemination via online chats, caregiver groups, and animal welfare organizations. The announcements will include a brief description of the study and an invitation for interested individuals to provide their telephone numbers. Those who express interest were contacted by phone to confirm eligibility and complete the survey. This recruitment strategy aims to maximize accessibility and ensure the inclusion of individuals actively engaged in feral cat caregiving. The structured questionnaire will assess compassion satisfaction, resilience, spiritual well-being, perceived social support, and Secondary Traumatic Stress component of Compassion Fatigue. This method ensures consistency in data collection and facilitates participation by caregivers regardless of geographic location. The data collection instrument consists of a structured questionnaire developed for this study. It includes sociodemographic variables (age, sex, main professional activity, and educational background) and information related to the feral cat colonies (location, number of animals, and whether the colony has been identified or legalized by the municipality).

Secondary Traumatic Stress component of Compassion Fatigue and Compassion satisfaction were assessed using the relevant subscales of the Professional Quality of Life Scale – Version 5 (PROQOL-5) [1, 25]. This instrument contains 30 items rated on a 5-point Likert scale (1 = “never” to 5 = “very often”). Scores are calculated by summing the respective 10 items for each subscale: items 18, 12, 34, 3, 20, 6, 22, 30, 16, and 27 for compassion satisfaction; and items 13, 9, 11, 25, 23, 7, 5, 2, 28, and 14 for Secondary Traumatic Stress component of Compassion Fatigue. Participants were asked to respond in reference to their experiences caring for feral cats. The Portuguese version of the PROQOL-5 was culturally adapted and validated by Carvalho [24] and later also by Duarte [26], showing good psychometric properties.

Spiritual well-being was measured using the Portuguese validate version [23] of the Spiritual Well-being Questionnaire (SWBQp) [27]. Scores are calculated by summing the respective 5 items for each subscale: items 5, 9, 14, 16 and 18 for Personal dimension; items 1, 3, 8, 17 and 18 for Community dimension; items 4, 7, 10, 12 and 20 for Environmental dimension and items 2, 6, 11, 13 and 15 for Transcendental dimension. Each dimension ranges from 5 to 25, with higher scores indicating higher levels of spiritual well-being in the respective dimension [23].

Resilience was assessed using the Portuguese version [22] of the original Resilience Scale by Wagnild and Young [28], which consists of 23 items grouped into four factors: (I) Perseverance, (II) Sense of life, (III) Serenity, and (IV) Self-reliance and self-confidence.

Perceived social support was evaluated using the Multidimensional Scale of Perceived Social Support [29], which includes 12 items covering support from family, friends, and significant others. Items are rated on a 7-point Likert scale (1 = “strongly disagree” to 7 = “strongly agree”), and the total score is obtained by summing all items, with higher scores indicating greater perceived social support. The Portuguese version of this scale [30] has demonstrated a stable three-factor structure and good psychometric properties.

The questionnaire included an open-ended question inviting participants to describe their main concerns regarding feral cats.

The study protocol was approved by the Ethics Committee of the Faculty of Medicine of the University of Porto (reference number: 332/CEFMUP/2025). All participants provided informed consent prior to participation. Participation was voluntary and participants could withdraw at any time without consequences. Data were collected through structured telephone interviews and were pseudonymized prior to analysis. All data were stored on password-protected and encrypted devices in accordance with the General Data Protection Regulation. The study involved minimal risk and no identifying information is reported.

The estimated minimum sample size was 100 participants. This sample size was determined through power analysis, assuming a statistical power of 0.80, a significance level of 5%, seven predictor variables, and a medium effect size (f^2^ = 0.15). Following dissemination of the project, a large number of volunteers expressed interest in participating, and all individuals who wished to take part and met the inclusion criteria were included, resulting in a final sample size exceeding the initially estimated minimum.

Initially, univariable linear regression analyses were performed to explore the association between each independent variable and the outcome. Variables with p<0.20 in univariable analysis were considered candidates for inclusion in the multivariable linear regression model, together with variables selected a priori based on theoretical relevance. Multicollinearity was assessed using variance inflation factors (VIF). Model assumptions were evaluated by inspection of residual plots, including assessment of linearity, normality of residuals (Q–Q plots), and homoscedasticity. Statistical significance was set at α = 0.05. All analyses were performed using R software.

## Results

A total of 294 individuals provided their contact information and expressed willingness to participate in the study. Of these, interviews were not conducted with 120 individuals because they did not meet the inclusion criteria, provided invalid mobile phone numbers, or did not answer the telephone after three attempts. In total, 174 interviews were conducted, of which two participants did not complete the questionnaire. Participants were mostly women, with a mean age of 53 years. The majority were employed and had completed higher education. Approximately 18% reported providing informal care to a dependent person. The number of cats cared for per colony varied widely. Most caregivers reported caring for between 5 and 14 cats (n=65), followed by 14–24 cats (n=39) and 25–34 cats (n=26). Smaller colonies with fewer than 5 cats were reported by 14 participants, while 11 cared for 35–44 cats and 19 reported caring for 45 or more cats. Because some categories had small sample sizes, the number of cats cared for was recorded into two groups (≤25 vs >25 cats), representing lower versus higher caregiving burden, grouping the three lowest and the three highest original colony-size categories.

Internal consistency was acceptable for the Compassion Satisfaction (Cronbach’s α = 0.72) and good for Secondary Traumatic Stress component of Compassion Fatigue (Cronbach’s α = 0.83). For the Spiritual Well-Being scale, Cronbach’s alpha coefficients were 0.80 (Personal), 0.66 (Community), 0.86 (Environmental), and 0.90 (Transcendental).In the resilience scale, Cronbach’s alpha coefficients were 0.77 for Perseverance, 0.75 for Meaning in Life, 0.68 for Serenity, and 0.50 for Self-Reliance and Self-Confidence. Importantly, the dimensions with lower internal consistency in our sample correspond to those that also presented lower Cronbach’s alpha values in the Portuguese validation study [22]. The relatively lower internal consistency of the Self-Reliance and Self-Confidence dimension should be taken into account when interpreting the results. Considering the social support scale, Cronbach’s α was 0.96 for the family dimension, 0.93 for the friends dimension, and 0.88 for the “significant others” dimension.

Considering, Secondary Traumatic Stress component of Compassion Fatigue, moderate levels was found in 47% of participants, whereas only 10% showed high levels. Overall, 150 participants (85%) showed high compassion satisfaction, whereas the remaining participants had moderate levels. In addition, the majority exhibited high levels of resilience. Regarding resilience dimensions, participants showed generally favorable levels, although with some variability across domains. Self-confidence presented the highest proportion of high scores, with approximately 58% of respondents classified in the high category, and only about 22% in the low category. Meaning in life also showed relatively positive results, with around 44% reporting high levels and roughly similar proportions in the medium and low categories (approximately 28% each). In contrast, serenity displayed a more balanced distribution, with a lower proportion of high scores (about 35%) and the highest proportion of low levels (approximately 37%) among the three dimensions.

Spiritual well-being subscale scores ranged from 5 to 25. The Personal and Community dimensions both showed a median score of 20 (interquartile range [IQR] = 5). The Environmental dimension presented the highest median score (24, IQR = 4). In contrast, the Transcendental dimension showed the lowest median score (median=18) and the greatest variability among participants (IQR = 11).

Most participants reported high perceived social support.

The distribution of respondents by district and the distribution of the Portuguese population by district according to the National Institute of Statistics are shown in Figure 1, suggesting a good geographic coverage of the sample.

**Figure 1.**
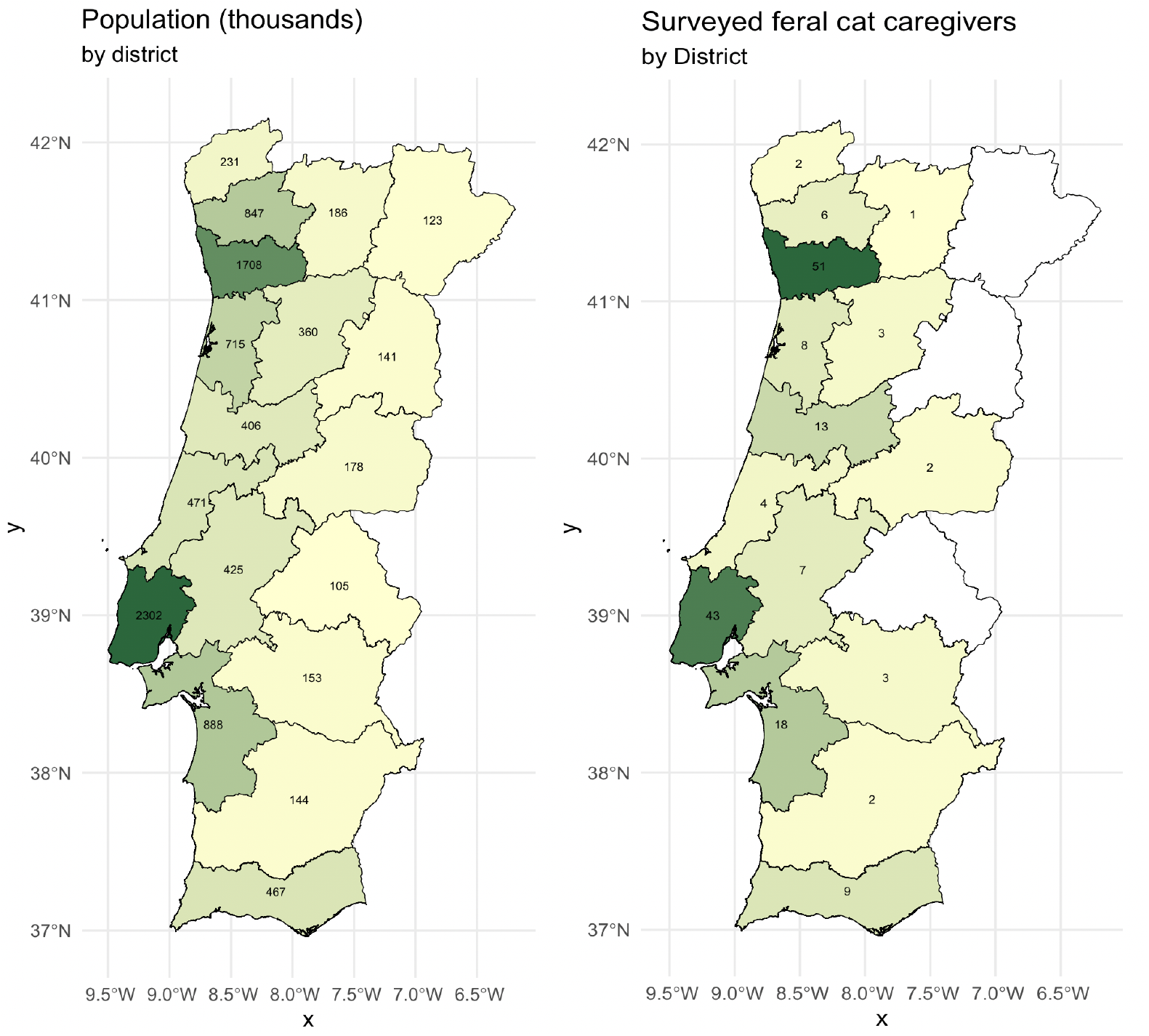
Geographic distribution by district of surveyed feral cat caregivers (right) and resident population according to the 2021 Portuguese Census (left). District population totals were obtained by aggregating municipality-level census data within each district. Two participants from the Autonomous Region of Madeira were not included in the map.

Table 1 presents the description of sociodemographic variables, characteristics of feral cat colonies and psychosocial measures of caregivers, along with the respective estimated coefficients (β) and p-values from univariate linear regression models, with Secondary Traumatic Stress as the dependent variable.

**Table 1.** Description and estimated coefficients (β) and p-values from univariate linear regression models, with Secondary Traumatic Stress component of Compassion Fatigue as the dependent variable.

| | description | $\beta$ | p |
| --- | --- | --- | --- |
| **Sociodemographic characteristics** |  |  |  |
| Age mean (sd) | 53 (11) | -0.191 | **<0.001** |
| Female gender n (%) | 164 (94) | 1.454 | 0.656 |
| Unemployed n (%) | 16 (9) | 10.302 | **<0.001** |
| Level of education: n (%) |  |  |  |
| Less than 12 years of schooling | 22 (13) | reference |  |
| Completed 12 years of schooling | 45(26) | 3.261 | 0.211 |
| Higher education | 107(61) | 2.690 | 0.251 |
| Informal caregiver for someone (human) n (%) | 31(18) | 0.044 | 0.982 |
| **Feral cat colonies characteristics** |  |  |  |
| Number of cats: n (%) |  |  |  |
| < 25 | 118 (68) | reference |  |
| 25 ou mais | 56 (32) | 5.500 | **<0.001** |
| Municipally registered colony: n (%) |  |  |  |
| No | 61 (35) | reference |  |
| Yes | 105 (60) | -1.796 | 0.265 |
| Dont't know | 80 (5) | 1.463 | 0.697 |
| Contact with any animal protection association: n (%) |  |  |  |
| No | 98 (56) | reference |  |
| Yes | 56 (32) | 0.520 | 0.757 |
| Be part of one | 20 (12) | 1.692 | 0.492 |
| Contact with the municipal veterinarian n (%) | 40 (23) | -1.000 | 0.579 |
| Contact with the parish council n (%) | 13 (8) | -1.747 | 0.545 |
| Cares for the cats alone n (%) | 113 (65) | 1.060 | 0.505 |
| Colony located near a high-traffic road n (%) | 97 (56) | -0.459 | 0.764 |
| **Psychosocial Measures** |  |  |  |
| Compassion Satisfaction median (IQR) | 47 (6) | -0.317 | 0.050 |
| Spiritual well-being: median (IQR) |  |  |  |
| Personal | 20 (5) | -0.742 | **<0.001** |
| Community | 20 (5) | -0.332 | 0.168 |
| Environmental | 24 (4) | 0.263 | 0.339 |
| Transcendental | 18 (11) | 0.062 | 0.617 |
| Resilience: median (IQR) |  |  |  |
| Perseverance | 6.0 (1.0) | -2.842 | **0.004** |
| Sense of life | 6.2 (1.0) | -2.269 | **0.016** |
| Serenity | 5.4 (1.1) | -2.923 | **<0.001** |
| Self-confidence | 6.2 (1.2) | 0.8511 | 0.374 |
| Perceived social support: median (IQR) |  |  |  |
| Family | 6.0 (3.0) | -1.347 | **0.001** |
| Friends | 6.0 (2.3) | -0.392 | 0.374 |
| Other relevant | 6.8 (1.0) | -0.406 | 0.499 |

The multivariable linear regression results are presented in Table 2. Higher Secondary Traumatic Stress component of was significantly associated with unemployment and with caring for larger cat colonies (≥25 cats), whereas increasing age was associated with lower Secondary Traumatic Stress levels. Among psychosocial variables, the resilience dimensions “sense of life” (positive association) and “serenity” (negative association) were significantly related to Secondary Traumatic Stress. In addition, greater perceived family social support was associated with lower fatigue. According to the standardized coefficients, unemployment showed the strongest association with Secondary Traumatic Stress.

**Table 2.** Description and estimated coefficients (β), standardized coefficients and p-values from multivariate linear regression model, with Secondary Traumatic Stress component of Compassion Fatigue as the dependent variable.

| | $\beta$ | Standardized<br>$\beta$ | p |
| --- | --- | --- | --- |
| **Sociodemographic characteristics** |  |  |  |
| Age mean (sd) | -0.155 | -0.175 | **0.019** |
| Unemployed n (%) | 10.399 | 0.302 | **<0.001** |
| **Feral cat colonies characteristics** |  |  |  |
| Number of cats: n (%) |  |  |  |
| < 25 |  | reference |  |
| 25 ou mais | 4.510 | 0.211 | **0.003** |
| **Psychosocial Measures** |  |  |  |
| Compassion Satisfaction median (IQR) | 0.068 | 0.032 | 0.682 |
| Spiritual well-being: median (IQR) |  |  |  |
| Personal | -0.460 | -0.163 | 0.076 |
| Community | 0.273 | 0.086 | 0.254 |
| Resilience: median (IQR) |  |  |  |
| Perseverance | -1.595 | -0.124 | 0.223 |
| Sense of life | 3.557 | 0.287 | **0.010** |
| Serenity | -2.407 | -0.263 | **0.005** |
| Perceived social support: median (IQR) |  |  |  |
| Family | -0.806 | -0.147 | **0.049** |

When asked about their main concerns regarding feral cats, the most frequently reported worries, mentioned by 32% and 31% of caregivers respectively, were the fear of dying, becoming ill, or having to relocate or change jobs, leaving no one to continue caring for the cats, and the lack of financial resources to care for the animals, including both feeding and veterinary treatment. Some caregivers also reported prioritizing the animals’ needs over their own, including, in some cases, forgoing their own medication in order to afford food and veterinary care for the cats. Subsequently, 27% of caregivers reported difficulties in treating the sick animals, due to challenges in recognizing illness, capturing the cats, or the cost of veterinary care. In addition, 23% expressed concern about discomfort experienced by cats living on the streets, and 22% feared intentional harm by people, such as poisoning or the destruction of shelters. Additionally, 15% of caregivers reported concern about the lack of understanding from the community and from authorities, such as municipalities and parish councils, regarding the caregivers’ work and the need to care for stray animals. Furthermore, 14% expressed concern about the sterilization of the cats in order to prevent colony overpopulation. Finally, 9% of caregivers reported concern about road accidents involving the cats, and 6% mentioned distress when a cat failed to appear on a given day, leaving them uncertain about its condition or whether it might be injured and in need of help. Only one caregiver expressed concern about municipal fines for feeding cats in the street, reporting that this had already occurred twice. Percentages may sum to more than 100%, as some caregivers reported multiple concerns.

## Discussion

The number of cats per colony was recorded in categories rather than as an exact count, given the inherent difficulty in estimating colony size and the frequent uncertainty reported by caregivers regarding the exact number of cats attending the colony. In our sample, 68% of the colonies comprised fewer than 25 cats, supporting the use of an average of approximately 20 cats per colony as a reasonable estimate. Using grouped category midpoints, the estimated mean colony size in our sample was approximately 20 cats. Combining this estimate with the reported national population of around 830,000 feral cats in Portugal [16], the number of caregiving colonies, and therefore informal caregivers, is likely to be in the order of several tens of thousands. Considering the assumptions and limitations of this approach, including uncertainty in colony size estimation, the possibility of multiple caregivers per colony, and the fact that not all feral cats may belong to identifiable colonies, this should be interpreted as a rough approximation rather than a precise countOur study shows that most participants exhibited moderate or high levels of Secondary Traumatic Stress component of Compassion Fatigue. Participants were predominantly women, with a mean age of 53 years; most were employed and had completed higher education. This profile contrasts with the common perception that feral cat caregivers are very elderly, socially isolated women with low educational attainment. Instead, our findings suggest a group of mainly middle-aged women who are generally active in daily life and well educated. A potential selection bias should, however, be considered. Older individuals and those with lower educational attainment may have been less likely to perceive the relevance of the study and therefore less likely to participate, which may have influenced the sample characteristics. Nevertheless, the results still identify a relevant subgroup of caregivers with this middle-aged, socially active, and well-educated profile.

Overall, most participants reported high compassion satisfaction, which is unsurprising given the sustained voluntary investment of time, effort, and even financial resources required for colony caregiving. However, compassion satisfaction was not significantly associated with lower Secondary Traumatic Stress after adjustment. This finding suggests that compassion satisfaction does not function as a protective factor against the Secondary Traumatic Stress component of Compassion Fatigue.

Most participants also reported high resilience and high perceived social support. Regarding spiritual well-being, the environmental dimension showed the highest scores, whereas the transcendental dimension showed the lowest. This pattern suggests that, in this population, spirituality may be more closely related to connection with nature and caregiving practices than to transcendental aspects.

The multivariable model suggests that Secondary Traumatic Stress in feral cat caregivers is influenced not only by external factors but also by the psychological meaning attributed to caregiving. As expected, caring for larger colonies (over 25 cats) and being unemployed were associated with higher Secondary Traumatic Stress, highlighting the importance of preventing vulnerable individuals from managing colonies alone and of providing adequate social and institutional support. Whereas older age appeared protective, possibly reflecting greater emotional regulation and experience in managing stressful situations.

Regarding resilience, the results indicate that its dimensions do not operate uniformly. Serenity was negatively associated with the Secondary Traumatic Stress, suggesting a protective effect. This dimension reflects the ability to accept adverse experiences and maintain emotional balance, which may help caregivers cope with chronic exposure to animal suffering and uncertainty. Caregivers with higher serenity may be better able to tolerate situations they cannot fully control, a common feature of feral cat management. In contrast, the sense of meaning in life dimension was positively associated with Secondary Traumatic Stress. This finding may indicate that caregivers who strongly perceive their caregiving role as central to their life purpose become more emotionally invested, and therefore more vulnerable to distress when faced with illness, death, abandonment, or limited capacity to help all animals. Rather than functioning as a buffer, meaning-driven involvement may intensify emotional exposure to suffering. In fact, one of the most frequently reported concerns among caregivers was the fear that, if they were to die or be forced to move, no one would continue caring for the cats, reflecting an underlying anxiety about having to abandon what they perceive as their responsibility or mission. These findings further highlight the importance of avoiding colonies being managed by a single individual and of promoting shared caregiving arrangements and municipality support. The Resilience Self-Reliance and Self-Confidence dimension, which showed less acceptable levels of internal consistency (Cronbach’s α = 0.50), was also the only resilience dimension not significantly associated with Secondary Traumatic Stress in the univariate analysis. The lower reliability of this subscale may have contributed to this finding. Nevertheless, the internal consistency of the remaining scales and dimensions used in this study was acceptable and generally good.

Perceived family social support showed a protective effect, reinforcing the idea that emotional sharing and validation outside the caregiving context may mitigate psychological strain; these findings further highlight the need for greater visibility, recognition, and external support for these caregivers.

Interestingly, spiritual well-being was not associated with Secondary Traumatic Stress in any dimension after adjustment. Although caregivers reported relatively high spiritual well-being, its influence appears to be expressed more in personal meaning and connection with nature than in buffering emotional exhaustion. This suggests that, in this population, coping with Secondary Traumatic Stress may depend more on emotional regulation and social support than on spiritual or transcendental resources.

A weakness of this study is that participants were recruited through online groups and animal protection associations, which may exclude feral cat colony caregivers outside these networks, potentially introducing a selection bias and limiting the generalizability of the results. However, recruitment required only the collection of a phone number followed by a telephone interview, which may have partially mitigated this limitation, as members of these groups could share contact details of caregivers not directly engaged online. Moreover, a good geographic distribution of participants across the country was achieved. Another weakness of this study is the potential presence of numerous confounding factors that we may not be able to identify or measure, a limitation inherent to any observational study of this nature. This highlights the inherent complexity of examining psychological distress in informal caregivers, as their experiences are shaped by a wide range of personal, environmental, and situational factors that may not be fully captured within the scope of this study.

A key strength of our study lies in its relevance, addressing a critical gap in understanding the mental health challenges faced by informal feral cat caregivers who often lack formal training or societal recognition, and it may contribute to the development of targeted training initiatives for caregivers. Another strength of this study is the use of scales validated for the Portuguese context, moreover, overall, the scales and subscales used demonstrated good reliability in our study, with the only concern being the lower internal consistency observed in the Self-Reliance and Self-Confidence dimension of the resilience scale. This enhances the robustness and credibility of the analyses by ensuring that the instruments have been culturally adapted and possess appropriate psychometric properties.

The study approaches health holistically, emphasizing the role of Secondary Traumatic Stress component of compassion satisfaction, resilience, spiritual well-being, and social support in mitigating psychological distress. By identifying the factors, both internal and external, that reduce psychological distress and protect against Secondary Traumatic Stress, the study highlights the need for targeted interventions to support the mental health and well-being of informal caregivers of feral cat colonies. By improving the well-being of caregivers, this study indirectly supports more effective care of feral cat colonies. This has broader public health implications, such as reducing disease transmission risks, improving sanitation, and addressing nuisance issues (e.g., noise and odors) in urban environments. Raising awareness about the challenges faced by informal caregivers can reduce stigma and promote community engagement, fostering healthier relationships between informal caregivers, the neighbors, and local authorities.

Interestingly, neither official colony registration nor caregivers’ contact with animal welfare organizations or the municipal veterinarian appeared to protect against Secondary Traumatic Stress component of, possibly reflecting the nature and scope of the support provided. Our findings support the need for clearer institutional responsibility and structured support for informal feral cat caregivers. In Portugal, municipalities are legally responsible for the management of stray animal populations, including population control and sterilization through TNR programs and basic sanitary supervision. However, ongoing feeding and routine veterinary care are often assumed in practice by informal caregivers, partly because municipal programmes frequently focus on sterilisation while not ensuring continuous feeding or clinical management, and partly due to caregivers’ reluctance to rely on municipal veterinary services, often related to concerns about the potential for inappropriate euthanasia decisions.

The association between higher Secondary Traumatic Stress and both larger colonies and unemployment suggests that financial burden plays a substantial role in caregivers’ psychological distress. In line with this, the costs of feeding and veterinary care were one of the concerns most frequently reported by participants. Significant expenses related to feeding and veterinary care, which, when combined with economic hardship, may considerably increase stress levels. Many feel morally unable to abandon their role due to compassion for the animals, yet struggle to bear costs. Therefore, municipalities, in coordination with parish councils and animal welfare organizations, should implement practical measures to facilitate the creation, monitoring, and support of managed colonies, while also ensuring ongoing veterinary care and population control. Frequently reported concerns were related to a perceived lack of understanding from the community and municipal authorities, as well as fear that people might harm the animals. These findings indicate the need for greater public education and awareness campaigns. At the same time, attention should be given to the protection and follow-up of informal caregivers, whose emotional burden represents a relevant public health concern. Addressing both animal welfare and caregiver well-being aligns with a One Health approach, recognizing the interconnected health of people, animals, and the community.

## Conclusion

This study shows that informal feral cat caregivers represent a sizable and largely unrecognized population, many of whom experience moderate to high levels of Secondary Traumatic Stress component of Compassion Fatigue. Fatigue was associated not only with external demands, particularly caring for larger colonies and economic vulnerability, but also with internal processes related to the meaning attributed to caregiving. Protective factors included older age, serenity, and perceived family support, suggesting that both emotional regulation and social support play an important role in coping. These findings indicate that psychological distress in this group results from the interaction between practical burdens and personal involvement in caregiving. Therefore, interventions should address both external needs, such as food for animals and veterinary support and coordinated collaboration with local authorities including greater public education and awareness campaigns, and internal coping resources, including psychosocial support and strategies that strengthen adaptive resilience. Supporting the psychological well-being of informal caregivers may therefore represent not only an individual-level intervention but also an integrated strategy to promote animal welfare, public health, and environmental quality in urban ecosystems.

## Data Availability

All data produced in the present study are available upon reasonable request to the authors

